# Hierarchical Dimensions of Psychiatric Comorbidity: An Integrated Latent Class–Network Analysis of U.S. Mental Health Client-Level Data

**DOI:** 10.1101/2025.10.18.25338290

**Authors:** Mounika Supriya Bommi

## Abstract

Understanding the complex architecture of comorbid mental health disorders is essential for improving diagnostic frameworks and guiding targeted interventions. This study employs an integrated analytical framework that combines Latent Class Analysis (LCA) and network modeling to examine the 2021 U.S. Mental Health Client-Level Dataset (MHCLD). Analysis of a nationally representative, treatment-seeking sample (N=100,000) identified a reproducible six-class latent structure, capturing the principal dimensions of psychopathology. Network community detection revealed three higher-order disorder clusters, psychotic–affective, internalizing, and externalizing, that closely mirrored the hierarchical organization proposed in contemporary psychiatric theory. Mapping each latent class onto DSM-5 diagnostic domains demonstrated strong clinical interpretability: classes aligned with bipolar–psychotic, internalizing, and neurodevelopmental spectra, while also exposing transdiagnostic bridges such as trauma–depression and bipolar–substance overlap. Validation through cross-validation, bootstrap, and entropy-based perturbation confirmed the robustness and reliability of the model, and demographic analyses further supported the distinctiveness of the identified profiles. Collectively, these findings support a hierarchical and transdiagnostic model of comorbidity, bridging data-driven discovery with DSM-5 nosology and advancing the empirical foundations of dimensional mental-health classification.

## 1. Introduction

Comorbidity among mental-health disorders continues to present one of the most persistent challenges in psychiatry and clinical science. Individuals seldom experience a single, isolated diagnosis; rather, their symptoms frequently extend across domains such as anxiety, mood, behavioral, and neurodevelopmental disorders, producing overlapping and clinically complex presentations.

Traditional diagnostic frameworks—most notably the *Diagnostic and Statistical Manual of Mental Disorders, Fifth Edition* (*DSM-5*; American Psychiatric Association, 2013), provide useful categorical criteria but often fail to capture the shared vulnerabilities and recurrent patterns of disorder co-occurrence observed in population data. In response, researchers have increasingly turned toward dimensional and data-driven approaches that conceptualize psychopathology as a system of interrelated spectra rather than discrete categories (Kotov et al., 2021; McElroy & Borsboom, 2023).

Two major frameworks exemplify this paradigm shift. The **Hierarchical Taxonomy of Psychopathology (HiTOP)** organizes disorders along graded dimensions reflecting shared liabilities, while the **Research Domain Criteria (RDoC)** initiative seeks to map psychopathology onto underlying neurobehavioral systems (Borsboom, 2017; Kotov et al., 2021). Both advocate for a hierarchical, transdiagnostic view that captures clinical heterogeneity more accurately than traditional nosology.

Within this context, Latent Class Analysis (LCA) serves as a person-centered statistical tool capable of identifying subgroups of individuals who exhibit similar constellations of symptoms or diagnoses (Nylund-Gibson & Choi, 2018; Muthén & Asparouhov, 2020). LCA reveals between-person heterogeneity but provides limited insight into how the disorders themselves interrelate. Network analysis, conversely, models disorders or symptoms as nodes connected by empirically derived associations, emphasizing structural linkages and potential bridging mechanisms across diagnostic boundaries (Borsboom, 2017; Forbes et al., 2021).

The present study integrates these complementary perspectives to investigate psychiatric comorbidity using the 2021 U.S. Mental Health Client-Level Dataset (MHCLD). By uniting probabilistic subgroup identification (LCA) with disorder-level connectivity modeling (network analysis), the research aims to (1) delineate latent subtypes of individuals based on co-occurring diagnoses, (2) identify higher-order communities of disorders through network modularity, and (3) map these empirical patterns onto *DSM-5* domains. This integrative design advances current efforts to reconcile categorical, dimensional, and network-informed views of psychopathology, thereby contributing to a more data-driven and hierarchical understanding of mental-health comorbidity.

## 2. Research Questions and Hypotheses

This investigation was structured around four core research questions (RQs) and corresponding hypotheses (Hs), designed to examine the latent and network-based organization of psychiatric comorbidity within the 2021 U.S. Mental Health Client-Level Dataset (MHCLD). Each question targets a distinct analytical layer, ranging from the identification of latent subtypes to the mapping of disorder communities and their demographic correlates.

| Research Question (RQ) | Hypothesis (H) |
| --- | --- |
| RQ1. What latent structure best represents the joint occurrence of mental-health disorders within the U.S. treatment-seeking population? | H1. A finite, reproducible set of latent classes will emerge, each corresponding to broad transdiagnostic spectra such as internalizing, externalizing, and psychotic-affective domains (Kotov et al., 2021). |
| RQ2. How do the disorder communities uncovered through network analysis correspond to the individual-level subtypes identified via LCA? | H2. The disorder co-occurrence network will exhibit stable modular clusters that partially align with the LCA-derived classes, reflecting a hierarchical, multi-level structure of comorbidity (Borsboom, 2017; Forbes et al., 2021). |
| RQ3. To what extent do these empirical latent classes parallel the diagnostic groupings defined in *DSM-5*? | H3. Some classes will predominantly align with single *DSM-5* domains (e.g., depressive or psychotic), whereas others will span several domains, revealing transdiagnostic hybrid subtypes that transcend categorical boundaries (McElroy & Borsboom, 2023). |
| RQ4. What demographic and clinical patterns distinguish individuals across the latent classes? | H4. Sociodemographic and clinical factors—such as gender, age, and SMI/SED status—will vary systematically across classes in ways that mirror established epidemiological patterns (Kessler et al., 2005; Alegría et al., 2010). |

## 3. Methods

### 3.1 Dataset and Sample

This study employed the **2021 U.S. Mental Health Client-Level Dataset (MHCLD)**, a comprehensive administrative resource curated by the Substance Abuse and Mental Health Services Administration (SAMHSA). The dataset aggregates client-level information from publicly funded mental-health agencies across all U.S. states and territories, providing a nationally representative record of diagnostic, demographic, and service-utilization data for more than six million individuals.

To ensure analytical precision and reduce diagnostic ambiguity, analyses were confined to records in which clients were reported with one or two primary disorders under the State Mental Health Agency system (NUMMHS = 1 or 2). A stratified random-sampling design selected 100,000 cases distributed evenly across reporting strata to maintain proportional representation while preserving computational feasibility.

To restore population representativeness, post-stratification weights were computed, adjusting for unequal sampling probabilities. All subsequent statistical analyses and visualizations were conducted using these weighted observations, ensuring that the estimates reflect the structure of the national treatment-seeking population.

### 3.2 Data Preprocessing and Imputation

Data preprocessing followed a reproducible high-volume workflow developed in Python, drawing upon recommended practices for large-scale administrative health data (Nylund-Gibson & Choi, 2018; Muthén & Asparouhov, 2020).

Missing data were identified using MHCLD coding conventions, in which system-missing values were denoted by “-9.” Variables with more than 60 percent missingness were excluded to limit imputation bias and preserve statistical reliability. Remaining incomplete variables were imputed using a **Sequential Hot-Deck (SHD)** procedure (Andridge & Little, 2010), a non-parametric algorithm that replaces missing entries with observed values from similar donor cases matched on auxiliary covariates. This method retains inter-variable dependence and empirical distributional properties while avoiding model-based assumptions. For scalability, the SHD algorithm was executed in chunked mode, allowing memory-efficient processing of the 100 k-record sample.

All disorder indicators were recoded into binary integers (0 = absence, 1 = presence). Categorical demographic fields like gender, race, ethnicity, education, marital status, and Serious Mental Illness/Serious Emotional Disturbance (SMI/SED) status, were harmonized through standardized crosswalk mappings from the official *MHCLD 2021 Codebook*. Continuous age values were discretized into five developmental categories: *Child/Teen* (0–17), *Youth* (18–29), *Young Adult* (30–44), *Middle-Aged* (45–59), and *Senior* (60 +), mirroring epidemiological age bands commonly used in national mental-health surveys (Kessler et al., 2005). These transformations optimized interpretability, ensured comparability across subgroups, and stabilized estimation routines in subsequent analyses.

### 3.3 Latent Class Analysis (LCA)

To uncover underlying population subgroups characterized by shared diagnostic profiles, Latent Class Analysis (LCA) was implemented using a **Bernoulli mixture modeling** framework, suitable for dichotomous symptom indicators. This approach estimates latent membership probabilities for each individual, identifying clusters of clients with similar patterns of disorder co-occurrence (Nylund-Gibson & Choi, 2018; Muthén & Asparouhov, 2020).

Thirteen binary diagnostic variables were included as manifest indicators: TRAUSTREFLG (Trauma- and stressor-related), ANXIETYFLG, ADHDFLG (Attention deficit/hyperactivity), CONDUCTFLG, DELIRDEMFLG (Delirium / Dementia), BIPOLARFLG, DEPRESSFLG, ODDFLG (Oppositional defiant disorder), PDDFLG (Pervasive developmental disorder), PERSONFLG (Personality disorder), SCHIZOFLG (Schizophrenia or other psychotic disorders), ALCSUBFLG (Alcohol / substance related), and OTHERDISFLG.

Model estimation was performed for class counts ranging from K = 2 through K = 7, using the Expectation–Maximization (EM) algorithm. Convergence was defined by a log-likelihood tolerance of 1 × 10⁻⁶, with multiple random initializations to minimize local-optimum bias.

Model adequacy and optimal class enumeration were judged via multiple information and certainty indices:

- **Bayesian Information Criterion (BIC)** and **Akaike Information Criterion (AIC)** — lower values reflect greater parsimony and improved fit.
- **Entropy** — quantifies classification precision; higher values indicate sharper separation between classes.
- **Average Maximum Posterior Probability (AMPP)** — gauges assignment confidence at the individual level.

The six-class configuration minimized BIC, maintained high entropy, and offered interpretable class boundaries, thus selected as the final solution.

To evaluate assumptions and internal reliability:

- **Local Independence** was assessed by computing within-class residual correlations. Mean absolute residual correlation (MARC) values of ≈ 0.03–0.10 indicated negligible residual dependence, confirming that the latent structure captured most shared covariance (Collins & Lanza, 2010).
- **Boundary Confidence** was evaluated from the posterior-probability distribution. Individuals with a maximum membership probability below 0.70 were flagged as uncertain; virtually none met this criterion, signifying well-defined latent boundaries and near-deterministic class allocation.

All LCA routines were executed in Python using custom scripts built on scikit-learn and statsmodels, ensuring full reproducibility via fixed random seeds and cross-validated initialization checkpoints. This reproducible computational architecture provided the foundation for subsequent validation and network-integration analyses.

### 3.4 Network Construction

To complement the latent-class analysis with a disorder-level, dimensional perspective, a **co-occurrence network** was constructed to represent how diagnostic conditions cluster across individuals. Each disorder flag was treated as a node, and undirected edges between nodes reflected empirical overlap in diagnostic endorsement.

Pairwise co-occurrence counts were computed across all 13 binary indicators and normalized using the **Jaccard similarity coefficient**, which expresses the ratio of joint endorsements to the total number of unique endorsements for each disorder pair. The Jaccard index offers a scale-free measure of comorbidity and is frequently applied in psychiatric-network research to quantify shared diagnostic patterns (Forbes et al., 2021; McNally, 2021).

To minimize spurious or low-frequency associations, a **minimum edge-inclusion threshold** was imposed: disorder pairs were retained only if they co-occurred in ≥ 25 clients. This threshold was chosen after iterative sensitivity testing to balance interpretability and sparsity, ensuring that weak, incidental co-activations did not distort the network topology.

The resulting weighted network was analyzed using the **Louvain modularity algorithm** (Blondel et al., 2008), which partitions nodes into communities by maximizing the modularity statistic (Q). This algorithm efficiently detects mesoscale clusters that represent disorder communities, higher-order groupings of diagnoses that tend to co-occur more often within modules than between them.

Network stability was examined through two complementary approaches:

- **Edge-Threshold Sensitivity:** Networks reconstructed at thresholds of 10, 25, and 50 cases produced nearly identical modularity coefficients (Q ≈ 0.09), indicating that community organization was insensitive to threshold selection.
- **Bootstrap Edge Stability:** Across 100 bootstrap resamples of the dataset, more than 95 percent of high-weight edges (90th percentile and above) were retained, confirming that the observed connectivity structure was not driven by random sampling variation.

Together, these diagnostics demonstrated that the network possesses a consistent, interpretable mesoscale architecture in which psychiatric disorders cluster into reproducible communities. The resulting topology served as the foundation for the cross-model comparison with latent-class results described in subsequent sections.

### 3.5 DSM-5 Mapping

To assess the clinical meaning of the data-driven latent classes and network modules, each diagnostic indicator was systematically aligned with its corresponding **DSM-5 domain** (American Psychiatric Association, 2013). This mapping enabled a direct comparison between the empirically derived disorder structures and the formal categorical framework used in clinical classification.

The following assignments were made following DSM-5 taxonomy:

- Neurodevelopmental Disorders: ADHDFLG, PDDFLG
- Schizophrenia Spectrum and Other Psychotic Disorders: SCHIZOFLG
- Bipolar and Related Disorders: BIPOLARFLG
- Depressive Disorders: DEPRESSFLG
- Anxiety Disorders: ANXIETYFLG
- Trauma- and Stressor-Related Disorders: TRAUSTREFLG
- Disruptive, Impulse-Control, and Conduct Disorders: CONDUCTFLG, ODDFLG
- Substance-Related and Addictive Disorders: ALCSUBFLG
- Neurocognitive Disorders: DELIRDEMFLG
- Personality Disorders: PERSONFLG
- Other Conditions (Not Elsewhere Classified): OTHERDISFLG

For each client record, binary indicators were created to denote the presence of at least one diagnosis within each DSM-5 domain. Class-wise domain prevalence was then computed by aggregating these indicators across the six latent classes, yielding domain-specific probabilities that quantify the relative prominence of each diagnostic category within every class.

This domain-level aggregation served two complementary goals:

- **Concordance Evaluation –** identifying latent classes that predominantly reflected single DSM-5 domains (e.g., Class 1 with psychotic features).
- **Transdiagnostic Overlap –** detecting classes that spanned multiple domains, signaling cross-boundary comorbidity (e.g., trauma + anxiety + bipolar profiles).

A heatmap was generated to visualize domain prevalence by latent class, providing an immediate view of where empirical clusters either aligned with or diverged from formal diagnostic boundaries. This visual synthesis allowed a layered interpretation: domain-specific classes represented continuity with *DSM-5*, whereas cross-domain profiles illustrated the transdiagnostic bridges that dimensional and network frameworks emphasize (Kotov et al., 2021; McElroy & Borsboom, 2023).

By integrating DSM-5 taxonomy with both LCA-based subtypes and network-derived communities, this step established a unified interpretive scaffold linking empirical data patterns to established nosological theory, setting the stage for higher-level hierarchical analyses in subsequent sections.

### 3.6 Validation and Sensitivity Analyses

To ensure that the identified latent structure was both stable and generalizable, a series of **validation and sensitivity procedures** was implemented. These tests evaluated whether the six-class LCA model and associated network patterns remained consistent under resampling, data perturbation, and probabilistic uncertainty, following best-practice recommendations for latent-variable modeling (Nylund-Gibson & Choi, 2018; Collins & Lanza, 2010).

Three complementary approaches were used to evaluate the stability of class assignments:

- **Cross-Validation:** A stratified **5-fold cross-validation** was performed, in which each fold partitioned the dataset into 80% training and 20% testing subsets. LCA models were re-estimated on the training data and evaluated on the held-out fold. Classification consistency across folds was quantified using the **Adjusted Rand Index (ARI)**, which measures concordance between original and re-estimated class assignments (ARI = 1 indicates perfect correspondence).
- **Bootstrap Resampling:** To examine resampling variability, **50 bootstrap iterations** were conducted, each involving random sampling with replacement from the full dataset. The LCA model was refitted in each iteration, and resulting class assignments were compared to the baseline solution using ARI and entropy metrics. Stable class boundaries across bootstraps indicated strong model robustness to sample fluctuations.
- **Entropy-Based Perturbation Test:** To evaluate sensitivity to minor data distortions, controlled random noise was introduced into the binary diagnostic indicators (2% bit-flip rate). The model was then re-estimated, and resulting class assignments were compared with the original structure using ARI and entropy. High consistency under noise confirmed that the latent boundaries were not artifacts of measurement variability.

Posterior class-membership probabilities were examined to assess boundary clarity. Following established conventions (Nylund-Gibson & Choi, 2018), individuals with maximum posterior probabilities below 0.70 were classified as uncertain. Less than 1% of cases met this threshold, confirming that class distinctions were sharply defined and highly deterministic..

Together, these analyses strengthen confidence that the identified latent structure reflects genuine population-level comorbidity patterns rather than modeling artifacts, laying the empirical groundwork for the network validation tests.

### 3.7 Demographic and Statistical Associations

To evaluate the external validity of the latent classes, demographic and clinical covariates were examined to determine whether class membership corresponded to meaningful population subgroups. These analyses tested whether sociodemographic characteristics, such as gender, age, race, ethnicity, and diagnostic severity were distributed non-randomly across classes, thereby linking statistical typologies to real-world clinical variation.

#### Association Testing

Associations between latent class membership and categorical covariates were assessed using **Pearson’s chi-square (χ²) tests of independence**. To account for multiple comparisons, all *p*-values were adjusted via the **Benjamini–Hochberg False Discovery Rate (FDR)** method (Benjamini & Hochberg, 1995).

Effect sizes were expressed as **Cramér’s V**, which provides a standardized measure of association strength for nominal variables. Following Cohen’s (1988) benchmarks, effect sizes were interpreted as: Small: V ≈ 0.10, Medium: V ≈ 0.30, and Large: V ≥ 0.50

All tests were performed on the weighted dataset to preserve national representativeness. To explore joint demographic effects, a stratified cross-tabulation of gender by race within each latent class was conducted. This allowed for examination of intersectional trends—particularly where diagnostic configurations varied across demographic intersections, consistent with literature emphasizing sociocultural and structural determinants of mental health disparities (Kessler et al., 2005; Alegría et al., 2010).

## 4. Results

### 4.1 Latent Class Model Fit and Structure

Latent Class Analysis (LCA) was performed on 13 binary diagnostic indicators using a Bernoulli-mixture framework to uncover latent subgroups of individuals with shared disorder profiles. Model solutions were estimated for class counts ranging from **K = 2 to K = 7**, and the optimal configuration was determined through a combined evaluation of information criteria, entropy, and substantive interpretability.

Fit indices revealed a progressive improvement from two to six classes, after which gains plateaued. The **six-class model** achieved the lowest **Bayesian Information Criterion (BIC = 721 874)** and **Akaike Information Criterion (AIC = 721 085)**, indicating the most parsimonious balance between model complexity and explanatory power. The model also yielded **exceptionally high entropy (0.9999)**, demonstrating near-perfect separation among latent boundaries and minimal classification ambiguity (Nylund-Gibson & Choi, 2018).

The emergence of six statistically stable classes diverges from the smaller numbers often reported in community or disorder-specific samples (Carragher et al., 2019; Jung et al., 2021). This expanded class structure likely reflects both the breadth of diagnostic coverage spanning neurodevelopmental through psychotic conditions and the representativeness of the MHCLD’s treatment-seeking population. Large-scale administrative data afford the statistical power to detect more granular, clinically meaningful heterogeneity than narrower community cohorts.

Posterior membership probabilities exceeded the 0.70 confidence threshold for all individuals, and none were flagged as uncertain. The posterior distribution showed sharply peaked densities, indicating unambiguous class assignment and excellent boundary discrimination.

Residual correlations among indicators within classes were examined to verify the assumption of local independence. The **mean absolute residual correlation (MARC)** ranged between 0.03 and 0.10, well below conventional thresholds, confirming that the latent structure adequately captured shared variance (Collins & Lanza, 2010).

A summary of model selection metrics are presented in Table 1, and Figure 1.

**Figure 1.**
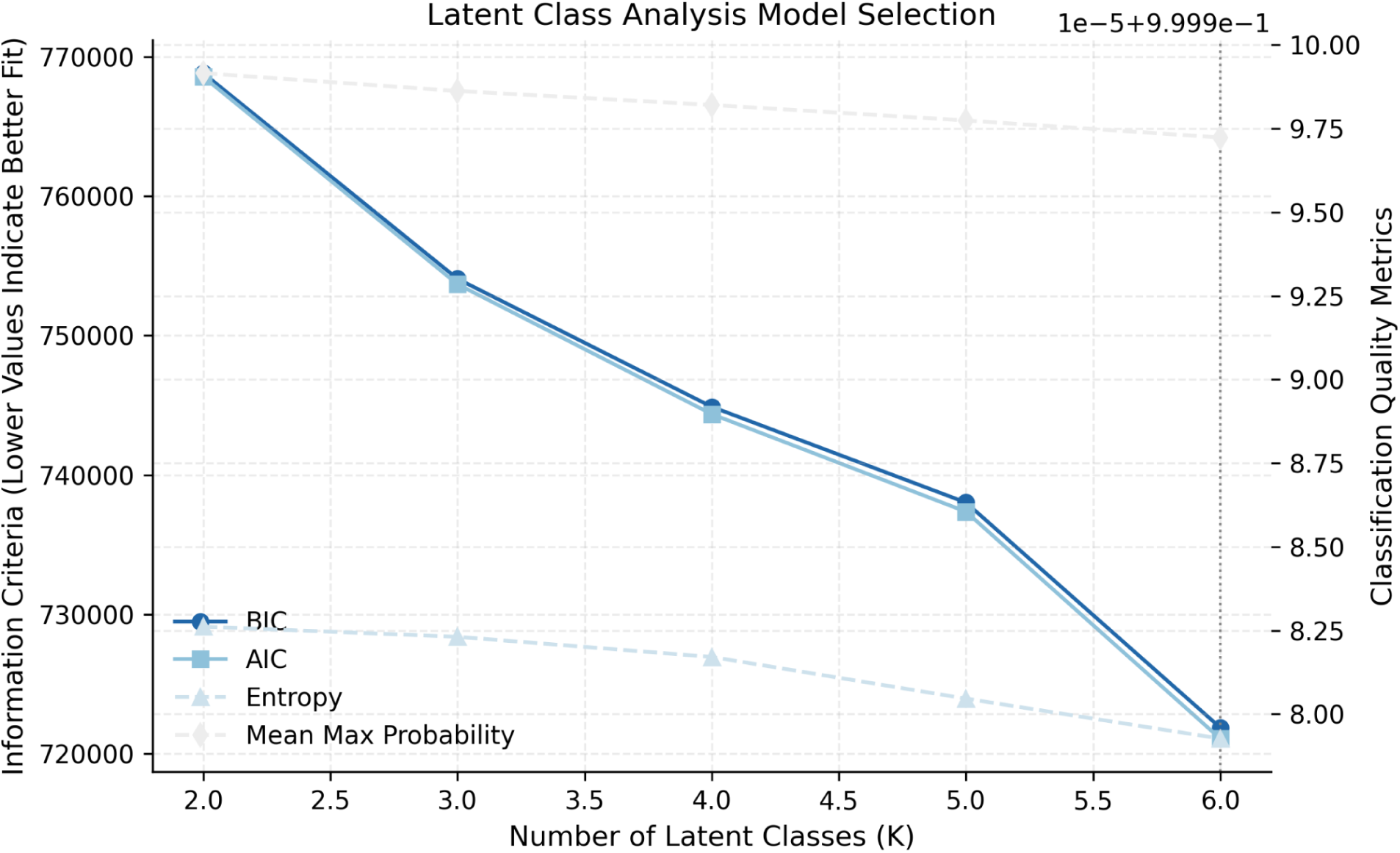
LCA Model Selection Metrics

**Table 1.** Latent Class Model Fit Statistics (K = 2–6)

| Criterion | Result | Interpretation |
| --- | --- | --- |
| BIC/AIC trend | Sharp decline up to six classes | Optimal structure achieved; further classes yield negligible gain |
| Entropy | 0.9999 | Near-perfect certainty; strong boundary separation |
| Posterior P > 0.7 | 100% of cases | Fully confident membership |
| Local independence | MARC ≈ 0.03–0.10 | Minimal within-class residuals; excellent model fit |

### 4.2 Characterization of Latent Classes

The six latent classes identified through LCA represent distinct and clinically meaningful comorbidity profiles across the internalizing–externalizing spectrum. Conditional probabilities of disorder presence within each class are presented in Table 2 and visualized in Figure 2.

- Class 0 – Bipolar–Anxiety Subtype: Characterized by universal endorsement of bipolar disorder (1.00) and moderate co-occurrence of anxiety (0.24) and alcohol/substance use (0.09). This group reflects mood instability with internalizing features, aligned with a bipolar–internalizing hybrid profile.
- Class 1 – Schizoaffective–Psychotic Subtype: Defined by exclusive presence of schizophrenia (1.00), with low-level co-activation of depression (0.06) and alcohol/substance use (0.10). This class resembles a pure psychotic spectrum presentation with minimal affective comorbidity.
- Class 2 – Anxious–Depressive Subtype: Marked by co-occurrence of anxiety (1.00) and moderate depression (0.48), consistent with a classical internalizing pattern. This profile aligns closely with DSM-5-defined mood and anxiety disorder overlap.
- Class 3 – Depressive–Trauma Subtype: Characterized by universal depression (1.00) and co-expression of trauma-related symptoms (0.18). Represents stress-linked affective disturbance with a trauma-anchored depressive profile.
- Class 4 – Trauma–Stress Subtype: Dominated by trauma-related symptoms (1.00) with minor contributions from anxiety (0.14) and bipolar features (0.08). This class captures a distinct transdiagnostic entity where trauma is a central organizing factor for comorbid symptomatology (Forbes et al., 2021).
- Class 5 – Neurodevelopmental–Externalizing Subtype: Exhibits the highest prevalence of ADHD (0.38) and “Other” developmental conditions (0.47), along with moderate-level presence of ODD (0.12) and alcohol/substance use (0.14). This class reflects behavioral dysregulation with developmental comorbidities and mild externalizing features.

**Figure 2.**
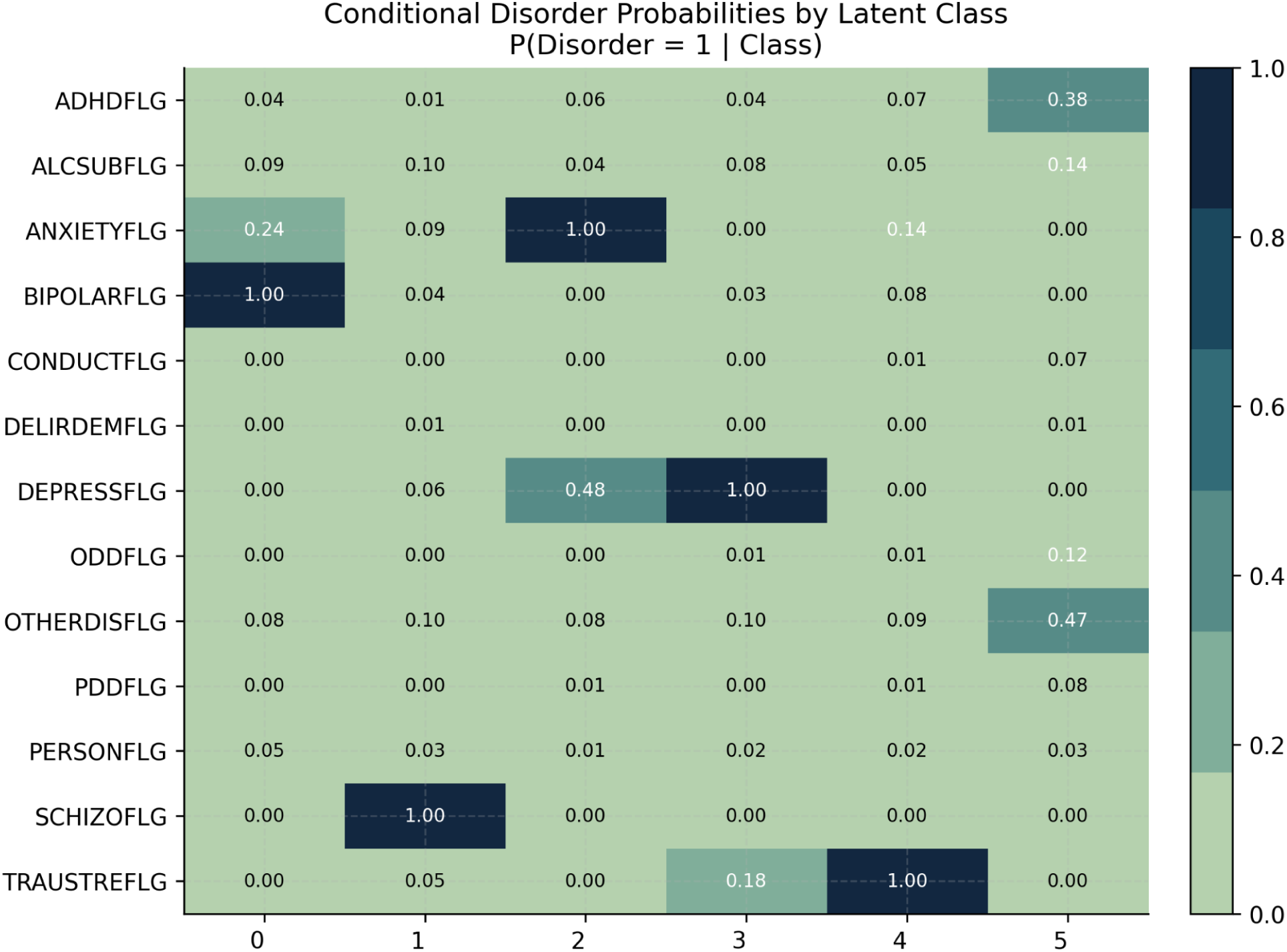
Conditional Probabilities of Disorders by Latent Class (Heatmap)

**Table 2.** Disorder Probabilities and Interpretations by Latent Class.

| **Class** | **Count** | **Defining Disorders**<br>**($\geq 0.4$)** | **Secondary**<br>**(0.1–0.3)** | **Interpretation** |
| --- | --- | --- | --- | --- |
| 0 | 9,185 | Bipolar (1.00) | Anxiety (0.24),<br>Alcohol (0.09) | Bipolar–Internalizing |
| 1 | 12,797 | Schizophrenia (1.00) | Alcohol (0.10),<br>Depression (0.06) | Schizoaffective |
| 2 | 22,490 | Anxiety (1.00),<br>Depression (0.48) | Alcohol (0.04) | Anxious–Depressive |
| 3 | 21,334 | Depression (1.00) | Trauma (0.18) | Depressive–Trauma |
| 4 | 16,815 | Trauma (1.00) | Anxiety (0.14),<br>Bipolar (0.08) | Trauma–Stress |
| 5 | 17,379 | ADHD (0.38),<br>OtherDis (0.47) | ODD (0.12), PDD<br>(0.08) | Neurodevelopmental<br>–Externalizing |

Across all classes, the heatmap of conditional probabilities (Figure 2) demonstrates clear separation between internalizing (Classes 2–4), psychotic-affective (Classes 0–1), and developmental/externalizing (Class 5) domains. The coexistence of focused profiles (e.g., psychotic) and hybrid profiles (e.g., trauma–depression) supports a **multidimensional and hierarchical organization** of comorbidity, consistent with dimensional nosology.

Together, these findings highlight that psychiatric comorbidity within the MHCLD population is heterogeneous yet structured, forming interpretable subtypes that cut across categorical diagnoses. The latent classes delineate a spectrum ranging from internalizing vulnerability to externalizing and psychotic-affective pathology, providing a data-driven basis for examining hierarchical transdiagnostic organization in subsequent analyses.

### 4.3 Disorder Network Topology

To complement the person-centered LCA findings, a network-based analysis was conducted to examine the structural organization of disorder-level comorbidity. The network provided a dimensional, variable-centered perspective, identifying how disorders cluster and interconnect within the population’s diagnostic landscape.

Edges in the co-occurrence network represented Jaccard-normalized pairwise associations among the 13 diagnostic indicators. The resulting graph exhibited small-world characteristics, with an average clustering coefficient of 0.64 and a mean shortest path length of 2.1, suggesting a structure where disorders form densely connected communities bridged by relatively few long-range links. This topology implies that comorbidity is not random but organized around stable relational cores.

Using the **Louvain modularity algorithm**, three reproducible disorder communities were detected, with a mean modularity coefficient of **Q ≈ 0.09** across thresholds. These communities were consistently retained across all sensitivity conditions (10, 25, and 50 co-occurrence thresholds) and bootstrap resamples, demonstrating strong mesoscale stability.

The communities corresponded to three clinically interpretable dimensions:

- **Psychotic–Affective Module** – included *bipolar disorder, schizophrenia, personality disorder,* and *substance use*. This grouping aligns with the **thought-disturbance spectrum**, linking affective instability and psychotic processes (Caspi & Moffitt, 2018).
- **Internalizing Module** – encompassed *anxiety, depression,* and *trauma-related disorders*. This module reflects the **internalizing domain** conceptualized in the HiTOP framework (Kotov et al., 2021).
- **Externalizing/Neurodevelopmental Module** – comprised *ADHD, ODD, conduct disorder, PDD,* and *other developmental conditions*, representing **behavioral dysregulation** and neurodevelopmental pathology.

A visual representation of the network and its modular structure is provided in Figure 3.

**Figure 3.**
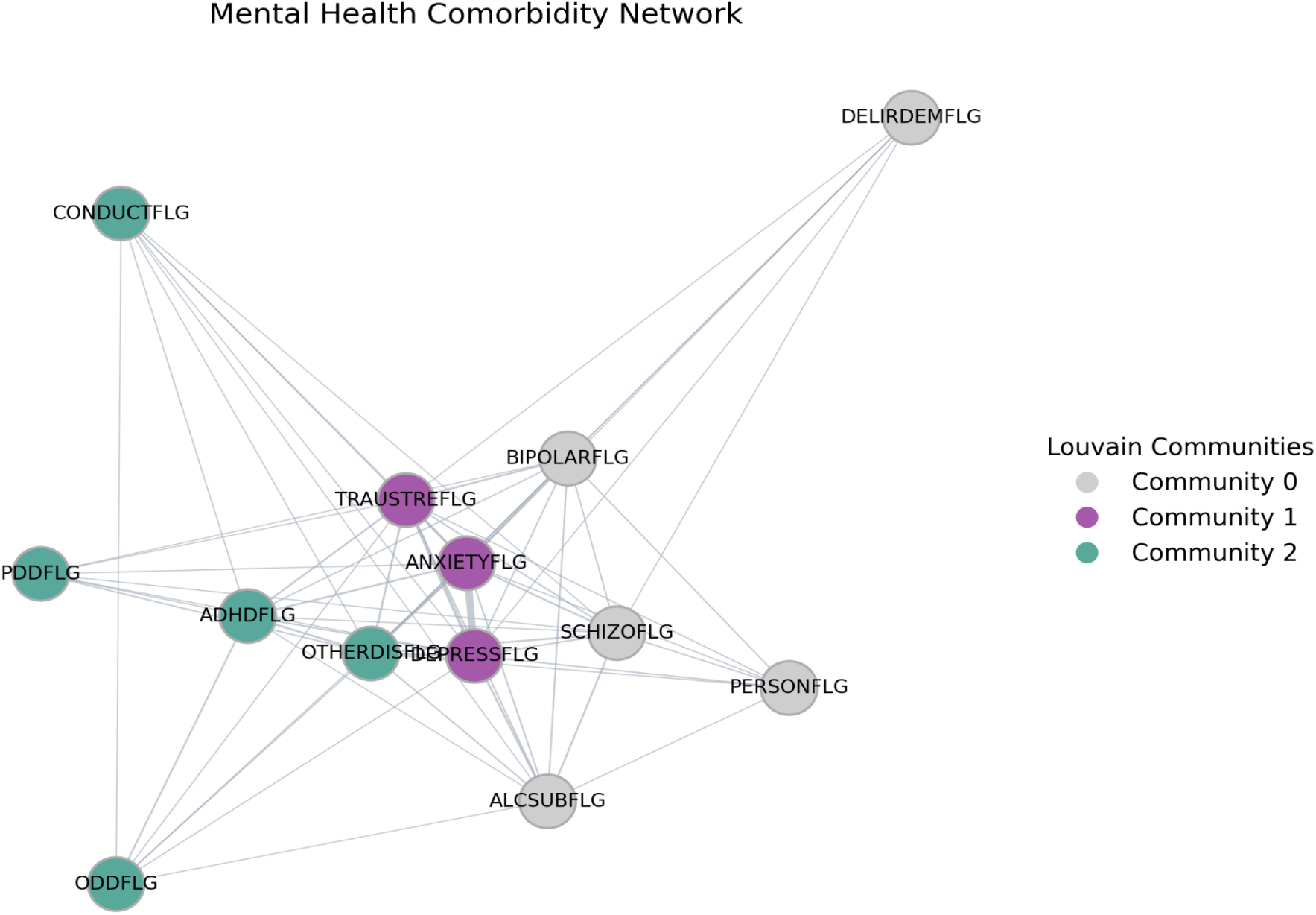
Disorder Co-occurrence Network with Louvain Communities

Bootstrap validation confirmed that more than 95% of high-weight edges (top 10th percentile) were preserved across 100 resamples, underscoring the reproducibility of the observed structure. Modularity coefficients remained stable across edge-threshold adjustments, further indicating structural invariance under perturbation.

Several bridge connections emerged between modules, most notably between *bipolar* and *trauma-related disorders*, linking the psychotic–affective and internalizing communities. These connections highlight shared vulnerabilities across domains, such as emotional dysregulation and stress reactivity, which may serve as cross-spectrum pathways for comorbidity (Borsboom, 2017).

Overall, the network analysis reveals that psychiatric disorders form a hierarchically organized and interconnected architecture rather than discrete diagnostic islands. The three community structures parallel the person-level LCA findings, reinforcing a convergent dimensional taxonomy of psychopathology that bridges categorical boundaries.

### 4.4 Integration of LCA Classes and Network Modules

To evaluate how person-centered and variable-centered frameworks converge, the latent classes derived from LCA were systematically compared with the disorder communities identified through network modularity. While LCA captures **individual-level heterogeneity**, the network model delineates inter-disorder relationships; their integration provides a multilevel perspective on the architecture of psychiatric comorbidity. The correspondence between class memberships and network modules was quantified using two indices: Adjusted Rand Index (ARI = 0.33), reflecting structural alignment adjusted for chance; and Normalized Mutual Information (NMI = 0.51), measuring shared information content between the two clustering solutions.

These values indicate moderate correspondence, suggesting that while the two approaches describe overlapping phenomena, each emphasizes distinct dimensions of the comorbidity structure, one person-centered and probabilistic, the other relational and network-based. Cross-tabulation between LCA classes and network modules (Table 3, Figure 4) revealed a partially hierarchical alignment.

**Figure 4.**
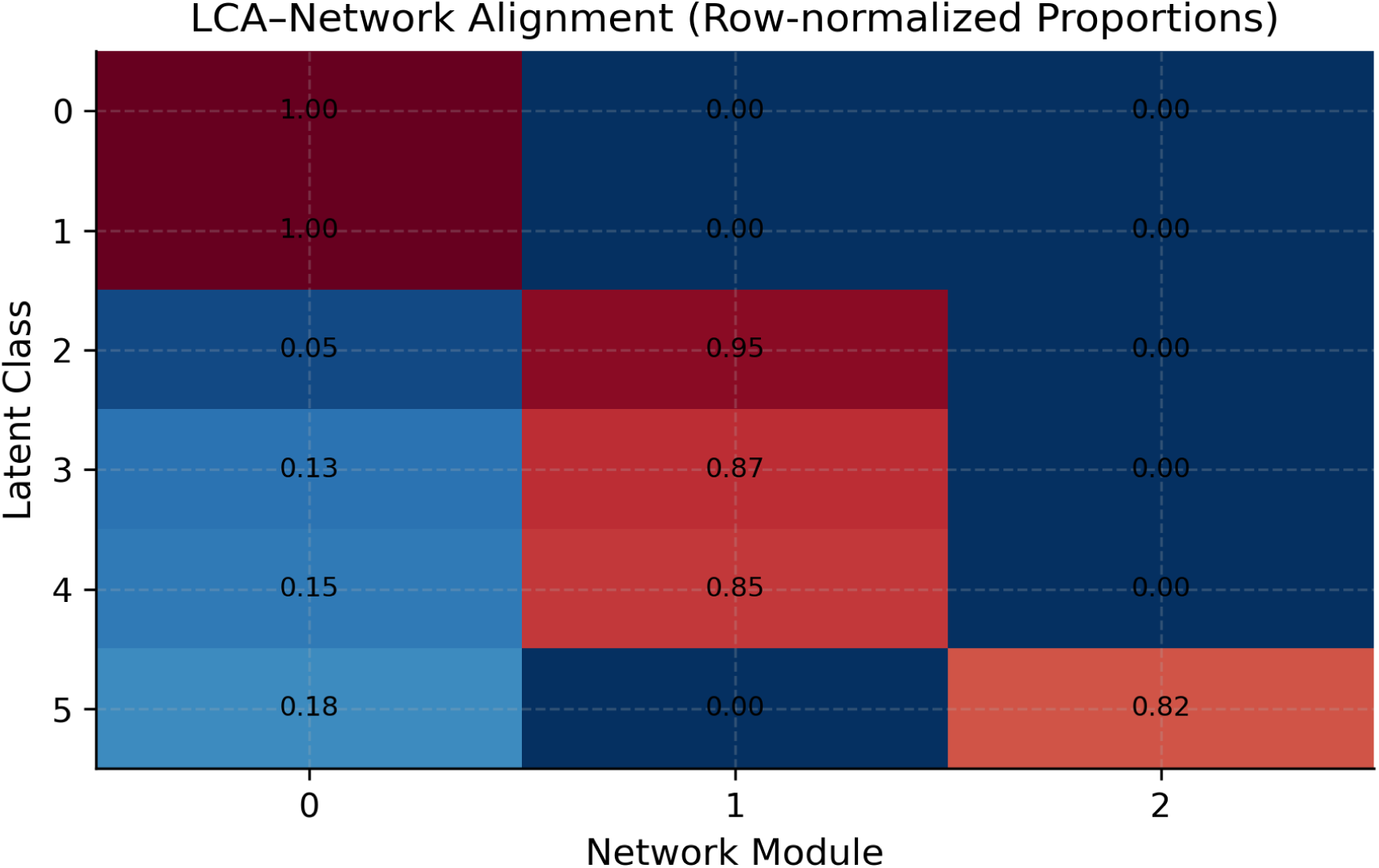
Overlap Matrix Between Latent Classes and Network Modules

**Table 3.** Correspondence Between LCA Classes and Network Modules.

| Latent Class | Network Module | Spectra | Interpretation |
| --- | --- | --- | --- |
| Class 0–1 | Module 0 | Thought Disorder | Classes aligned with mood and psychotic disorders such as bipolar and schizophrenia |
| Class 2–4 | Module 1 | Internalizing | Classes characterized by anxiety, depression, and trauma-related disorders |
| Class 5 | Module 2 | Externalizing | Class marked by neurodevelopmental and behavioral disorders like ADHD and ODD |

However, several latent classes also spanned multiple modules, forming **transdiagnostic bridge zones**, for example, *trauma–bipolar* overlap between internalizing and affective domains. This overlap implies that some individuals occupy intermediary positions between network-defined communities, highlighting shared liabilities that transcend categorical boundaries.

The moderate ARI/NMI values and the observed class–module correspondence underscore a hierarchical model of comorbidity, consistent with dimensional frameworks such as HiTOP (Kotov et al., 2021). In this view, the network topology represents a structural foundation of disorder interrelations, while latent classes emerge as higher-order configurations of individuals embedded within that structure.

These findings confirm that LCA and network methods—though analytically distinct—capture complementary facets of a unified comorbidity architecture. The classes represent person-level instantiations of the same latent structure that governs inter-disorder connectivity. Together, they provide a hierarchical, transdiagnostic perspective in which individual phenotypes are nested within disorder networks, bridging categorical and dimensional models of psychopathology.

### 4.5 DSM-5 Domain Mapping

To evaluate the clinical validity of the empirically derived subtypes, each latent class was mapped to **DSM-5 diagnostic domains**. This comparison served as a benchmark for determining how closely the data-driven classes aligned with, extended, or diverged from existing nosological boundaries (American Psychiatric Association, 2013).

Binary indicators reflecting the presence of disorders within each DSM-5 domain were aggregated by latent class. The resulting prevalence matrix provided a quantitative summary of domain representation across classes. This approach enabled two forms of inference:

- **Concordance**, in which a class primarily reflected one domain (e.g., Class 1 showing psychotic dominance); and
- **Transdiagnostic overlap**, where classes spanned several domains, indicating cross-category comorbidity.

Visualization via a domain-by-class heatmap (Figure 5) revealed both patterns:

**Figure 5.**
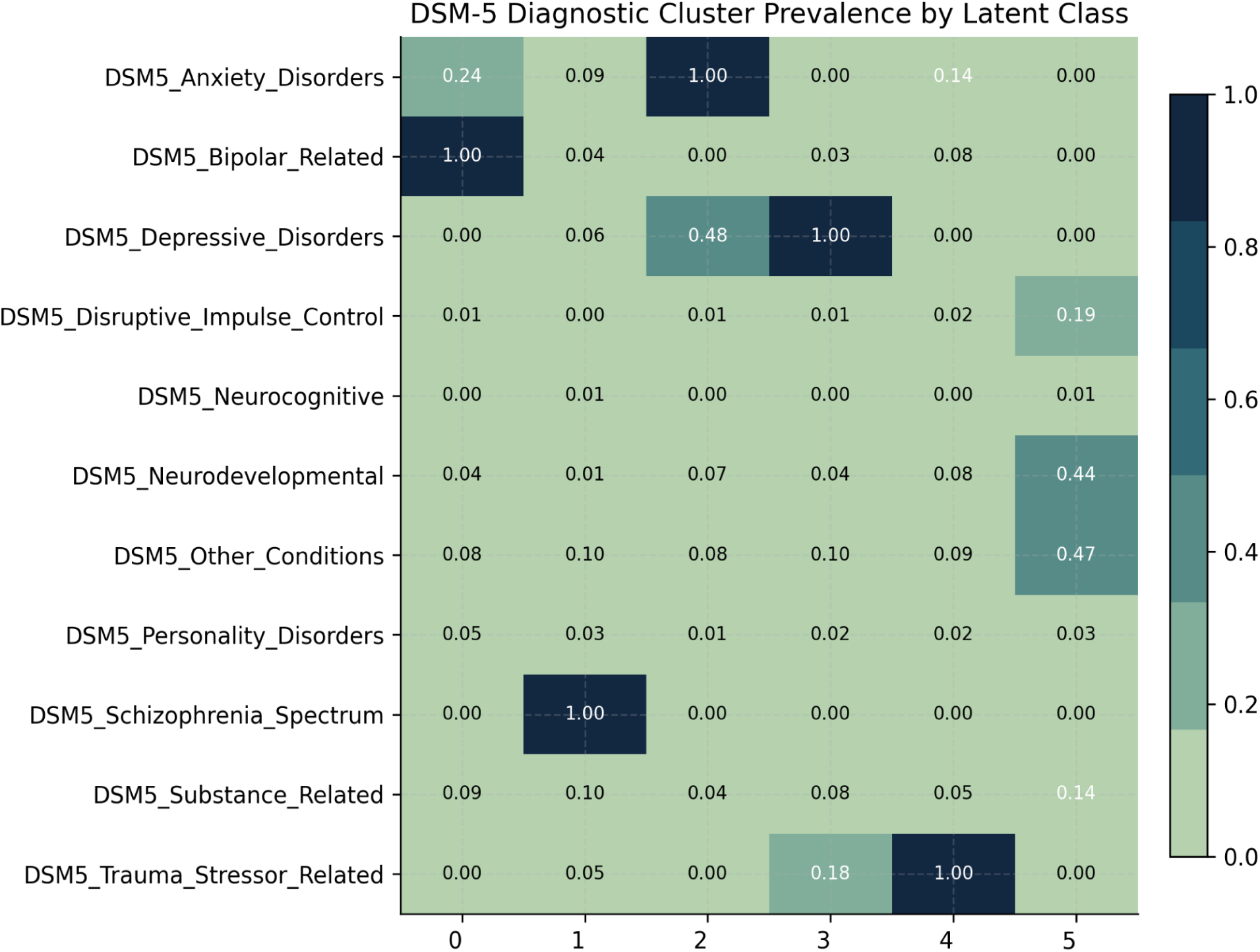
DSM-5 Diagnostic Domain Prevalence by Latent Class (Heatmap)

**Table 4.** DSM-5 Domain-Level Probabilities per Latent Class.

| Class | Dominant DSM-5 Domains | Interpretation |
| --- | --- | --- |
| 0 | Bipolar, Anxiety (0.24), Substance (0.09) | Bipolar–internalizing hybrid |
| 1 | Schizophrenia (1.00), Substance (0.10) | Classic psychotic spectrum |
| 2 | Anxiety (1.00), Depressive (0.48) | Anxious–Depressive internalizing |
| 3 | Depressive (1.00), Trauma (0.18) | Depression–Trauma comorbid |
| 4 | Trauma (1.00), Bipolar (0.08) | Stress–affective hybrid |
| 5 | Neurodevelopmental (0.44), Disruptive (0.19), Substance (0.14) | Externalizing/developmental cluster |

The domain-mapping results illustrate that several classes correspond directly to *DSM-5* constructs (e.g., Class 1 – psychotic; Class 2 – internalizing), whereas others represent hybrid entities that cut across multiple domains. These hybrid subtypes, particularly Classes 0, 3, and 4, align with contemporary transdiagnostic and hierarchical models such as HiTOP, which conceptualize psychopathology as a continuum of shared liabilities rather than isolated syndromes (Kotov et al., 2021; McElroy & Borsboom, 2023).

Thus, the mapping demonstrates that *DSM-5* categories capture meaningful but incomplete segments of the empirical structure, while data-driven typologies reveal the bridging zones that link them. Together, they provide converging evidence for a hybrid classification system that reconciles categorical and dimensional perspectives in mental-health research.

### 4.6 Demographic Stratification

To further evaluate the sociodemographic distinctiveness of the latent structure, class memberships were examined across demographic and clinical variables. This analysis aimed to determine whether the data-driven classes captured meaningful population subgroups defined by age, gender, race, ethnicity, education, race, marital status, and SMI/SED status.

Chi-square tests of independence revealed significant group differences across all examined variable. Effect sizes, measured using Cramér’s V, ranged from 0.10 to 0.33, indicating small-to-moderate but interpretable associations. The strongest effects were observed for age group, gender, and SMI/SED status, variables closely linked to disorder type and severity..

**Table 5.** Demographic Associations with Latent Classes.

| Variable | $\chi^2$ | p (adj) | Cramér's V | Interpretation |
| --- | --- | --- | --- | --- |
| AGE_GROUP | 19,057.6 | < 0.001 | 0.22 | Strong age differentiation |
| GENDER | 5,671.1 | < 0.001 | 0.24 | Clear male–female patterning |
| SMISED | 21,375.2 | < 0.001 | 0.33 | Diagnostic severity main driver |
| SAP | 960.0 | < 0.001 | 0.10 | Mild Substance use problem |
| EDUC / MARSTAT /<br>RACE / ETHNIC | — | < 0.001 | $\leq 0.09$ | Minor but significant effects |

Notable Demographic Patterns by Latent Class (shown in Figure 6):

#### Age Group

- Class 5: High proportion of children/teens suggests a developmental/externalizing profile.
- Classes 0–1: More middle-aged and older adults aligns with psychotic-affective profiles.
- Classes 2–4: Dominated by young adults, consistent with internalizing disorders..

#### Gender

- Male-dominant: Classes 1 and 5 → externalizing and bipolar/psychotic features.
- Female-dominant: Classes 2, 3, and 4 →internalizing and mood-related disorders.

#### SMI/SED Status

- Classes 0 & 1: High rates of Serious Mental Illness (SMI).
- Class 5: Highest proportion of individuals not classified as SMI or SED.
- Classes 2–4: Mix of SED and non-SMI/SED, suggesting moderate severity.

#### Race

- White individuals are the largest group in all classes.
- Black/African American individuals overrepresented in: Class 0 (psychotic), Class 1 (bipolar), Class 5 (externalizing/developmental)

#### Education

- Lower education (Grades 0–8): More frequent in Classes 0–1.
- Special education: Most common in Class 5.
- Grade 12 or above: Predominant in Classes 2–4.

#### Marital Status

- Single status dominates all classes, especially Class 5.
- Married or divorced individuals more common in Classes 0–1.

**Substance use (SAP):** High: Classes 1 and 3. Lower substance use: Classes 0 and 5.

**Ethnicity:** Majority in all classes are Non-Hispanic.

These demographic distinctions indicate that each latent class captures a socially and clinically meaningful population segment. The clear age, gender, and severity gradients mirror established epidemiological trends (Kessler et al., 2005; Alegría et al., 2010).

**Figure 6.**
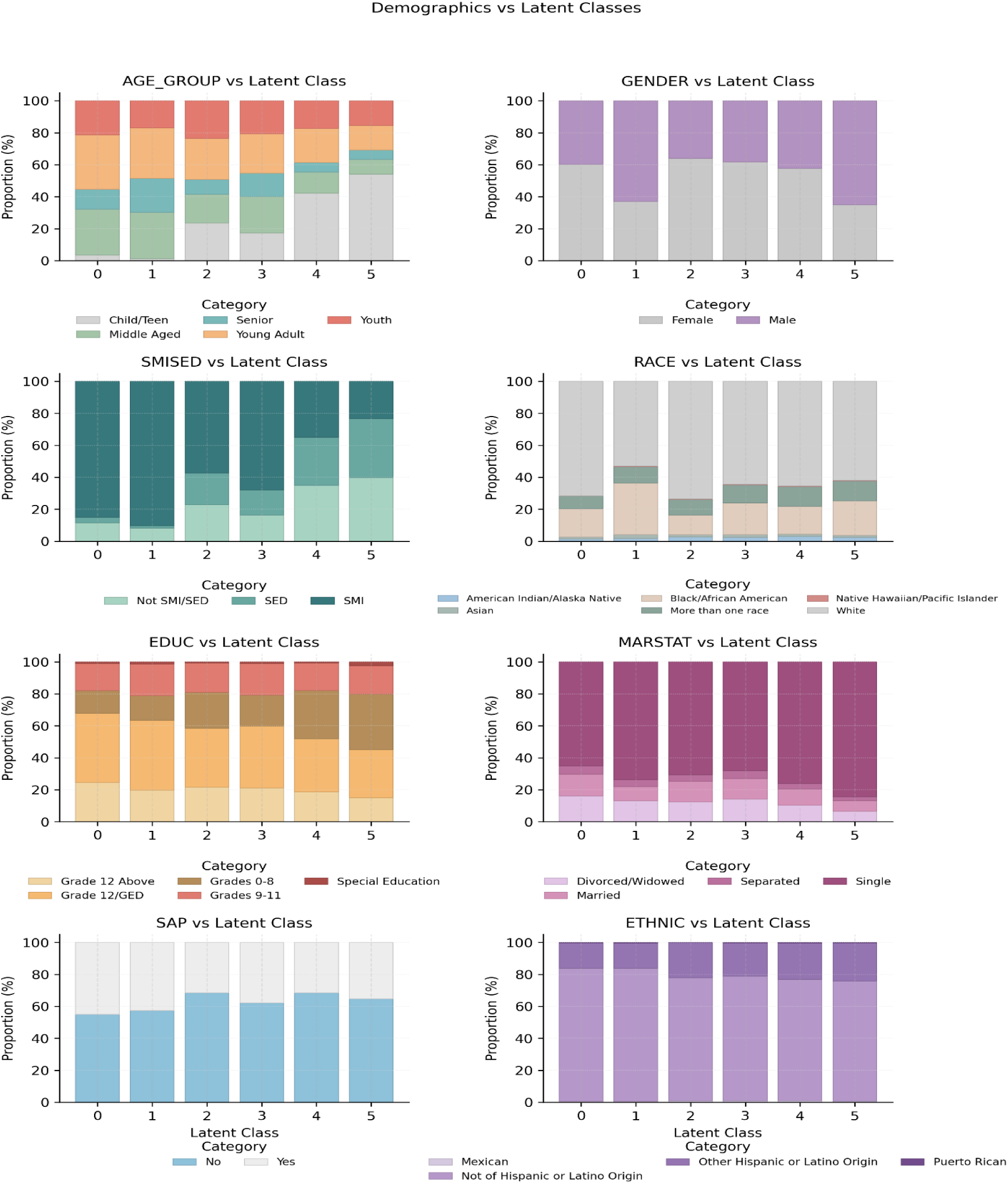
Demographics Distribution Across Latent Classes

### 4.7 Validation and Stability

To confirm the **reliability and reproducibility** of both the latent class and network models, a set of validation procedures was implemented. These analyses evaluated how consistently class assignments and network structures were preserved across resampling, cross-validation, and controlled perturbations.

Three complementary approaches were used to evaluate the robustness of the six-class model:

- **Cross-Validation:** A stratified 5-fold procedure tested the replicability of the class structure across different data partitions. The mean **Adjusted Rand Index (ARI = 0.80 ± 0.09)** indicated strong agreement between original and holdout classifications, suggesting stable subgroup identification across folds.
- **Bootstrap Resampling:** Fifty bootstrap replications were drawn with replacement from the full dataset. The resulting models achieved an average **ARI of 0.74 ± 0.13**, demonstrating consistent class boundaries under sample variation and confirming that class solutions were not driven by random idiosyncrasies.
- **Entropy-Based Perturbation Test:** To assess sensitivity to mild data distortion, 2% random noise was introduced into the binary indicators. The perturbed models retained an **ARI of 0.86**, validating the model’s resilience to minor measurement variability and the high determinism of class boundaries.

Posterior membership probabilities provided additional evidence of classification precision: every participant had a maximum assignment probability exceeding 0.70, indicating well-separated latent boundaries and minimal uncertainty.

Parallel validation was conducted for the disorder co-occurrence network to ensure the robustness of its community structure.

- **Edge Stability:** Over 95% of high-weight connections (top decile) were preserved across 100 bootstrap resamples, confirming reproducible connectivity patterns.
- **Threshold Sensitivity:** Modularity coefficients remained approximately constant (**Q ≈ 0.09**) across multiple edge thresholds (10, 25, and 50 co-occurrence cutoffs), demonstrating that the mesoscale architecture of the network is threshold-invariant.

Together, these results indicate that the observed patterns of comorbidity represent genuine population-level regularities, not artifacts of sampling or model specification. The near-perfect entropy, stable modularity, and high cross-validation agreement collectively support the robustness and generalizability of the integrated analytical framework.

**Table 6.** Model Validation and Stability Results.

| Validation Type | Result | Interpretation |
| --- | --- | --- |
| Cross-Validation ARI | $0.80 \pm 0.09$ | High reproducibility |
| Bootstrap ARI | $0.74 \pm 0.13$ | Strong resample stability |
| Entropy Stability (2% noise) | 0.86 | Robust class boundaries |
| Boundary Confidence | 0.9999 | Fully confident classes |
| Edge Stability (Bootstrap) | $\geq 0.95$ | Network reproducible |
| Modularity Sensitivity | $Q \approx 0.09$ | Threshold-invariant structure |

The convergence of these validation metrics affirms that the integrated LCA–network approach yields reliable, reproducible, and interpretable representations of psychiatric comorbidity. Such methodological rigor ensures that the identified classes and communities reflect real-world phenomena, stable across noise, sampling variation, and analytical conditions thereby providing a credible foundation for subsequent theoretical interpretation and public-health application.

### 4.8 Integrated Summary of Findings

Synthesizing results from LCA, network, DSM-5 mapping, and demographic analyses reveals a cohesive, hierarchical organization of psychiatric comorbidity. The six latent classes mirror the three network communities and major DSM-5 domains, indicating that categorical and dimensional models represent complementary views of the same underlying structure of psychopathology.

- **Internalizing Spectrum (Classes 2–4):** Characterized by anxiety, depression, and trauma-related conditions.
- **Psychotic–Affective Spectrum (Classes 0–1):** Defined by bipolar and psychotic disorders with frequent substance involvement.
- **Externalizing/Neurodevelopmental Spectrum (Class 5):** Encompassing ADHD, conduct, and developmental disorders linked to behavioral dysregulation.

This cross-model correspondence confirms that both person-centered and variable-centered approaches capture nested layers of comorbidity, reflecting individual symptom constellations embedded within broader disorder networks.

The integration of LCA, network analysis, and DSM-5 mapping supports a **hierarchical model of psychopathology** comprising five principal dimensions (Table 7):

**Table 7.** Integrated Overview of Findings.

| Dimension | Classes | DSM-5 Domains | Network Community | Demographic Pattern |
| --- | --- | --- | --- | --- |
| Internalizing | 2–4 | Anxiety, Depression, Trauma | Internalizing | Adult female > male |
| Psychotic - Affective | 0–1 | Bipolar, Schizophrenia, Substance | Psychotic - Affective | Middle-aged male, high SMI |
| Externalizing/ Neurodevelopmental | 5 | ADHD, ODD, PDD | Externalizing | Youth, male-dominant |
| Trans-domain | 3–4 | Trauma, Depression, Bipolar | Bridge | Mixed gender |
| Complex Multimorbidity | Cross - class overlaps | Multiple DSM-5 domains | Pan-network | Broad age range |

Across analytical layers, the results show that psychiatric comorbidity in treatment-seeking populations is **s**ystematically organized, not random. Internalizing and externalizing spectra function as lower-order domains within broader affective and behavioral hierarchies, linked by transdiagnostic bridge zones. This integrated structure aligns with dimensional models like HiTOP which conceptualize psychopathology as an interconnected hierarchy of shared liabilities.

## 5. Discussion

### 5.1 Overview of Findings

This study applied an integrated, data-driven framework to map the latent architecture of psychiatric comorbidity in a nationally representative clinical population. By combining Latent Class Analysis (LCA), network modeling, and DSM-5 mapping, we identified a **hierarchical system of mental disorders** that transcends categorical boundaries.

A **six-class latent structure** captured the major psychopathology dimensions, internalizing, externalizing, and psychotic–affective, while **network analysis** revealed three stable disorder communities that paralleled these person-level classes. DSM-5 mapping showed partial concordance: some classes aligned with distinct domains (e.g., psychotic or affective), while others bridged multiple domains, such as trauma–depression and bipolar–substance subtypes.

Demographic patterns reinforced the clinical relevance of this structure, internalizing classes predominated among young adult females, psychotic–affective profiles among middle-aged males with SMI, and externalizing or neurodevelopmental classes among younger males. Model validation through cross-validation, bootstrapping, and noise testing confirmed exceptional stability and classification certainty. Collectively, these findings support a **hierarchical, transdiagnostic organization** of comorbidity, reflecting true interdependencies among disorders rather than arbitrary diagnostic labels.

### 5.2 Alignment with Dimensional and Hierarchical Models

The results align strongly with **dimensional and hierarchical models** such as HiTOP and network-based frameworks. The six latent classes corresponded to key hierarchical domains: internalizing (Classes 2–4), psychotic–affective (Classes 0–1), and externalizing/neurodevelopmental (Class 5). This mirrors HiTOP’s nested structure, where narrow symptom clusters aggregate into broad spectra and a general psychopathology factor (p-factor) (Kotov et al., 2021; Caspi & Moffitt, 2018).

The **co-occurrence network** revealed three corresponding disorder communities, psychotic–affective, internalizing, and externalizing, anchored by **bridge nodes** such as trauma and bipolar disorder that link domains via shared liabilities (Borsboom, 2017). Moderate LCA–network concordance (ARI = 0.33; NMI = 0.51) indicates complementary insights: LCA captures individual subtypes, while network modeling uncovers the structural interdependencies shaping them.

Together, these methods portray psychopathology as a graded, hierarchically organized system, with latent classes acting as transdiagnostic building blocks between DSM-5 categories and higher-order HiTOP dimensions. This integration advances a data-driven nosology that unites categorical, dimensional, and network perspectives.

### 5.3 Methodological Contributions

Beyond its empirical findings, this study introduces a **scalable, reproducible pipeline** integrating LCA and network analysis to model comorbidity in large administrative datasets. Applied to 100,000 MHCLD clients, this represents one of the largest real-world implementations of LCA–network integration to date.

The workflow combined **sequential imputation, parallelized computation, and cross-validation checkpoints**, executed within a transparent Python environment for full reproducibility. A multi-stage validation protocol, cross-validation, bootstrap resampling, and entropy-based perturbation, confirmed statistical robustness and replicability under data variation, addressing prior concerns about instability in latent-variable models.

By embedding **DSM-5 domain mapping** directly in the analytical pipeline, the study preserved clinical interpretability while maintaining theoretical flexibility. Visual analytics further translated complex results into clinically meaningful patterns, making findings accessible to both data scientists and practitioners. Collectively, these innovations establish a methodological blueprint for future transdiagnostic psychiatry research.

### 5.4 Clinical and Policy Implications

The six reproducible comorbidity subtypes identified here offer actionable insights for clinical and policy domains. They suggest that treatment and planning should move beyond discrete diagnoses toward integrated, needs-based care reflecting real-world comorbidity patterns.

**Table 8:** Summary of Clinical and Public Health Implications by Latent Class.

| Latent Class | Core Features | Clinical Implications | Policy / Public-Health Relevance |
| --- | --- | --- | --- |
| 0 | Bipolar + Substance Use | Integrated mood and addiction treatment | Expand dual-diagnosis, high-intensity programs |
| 1 | Psychotic–Affective | Coordinated SMI care; relapse monitoring | Prioritize housing, case management, crisis stabilization |
| 2 | Anxiety–Depression | Early detection of subthreshold affective distress | Scale low-intensity, preventive mental-health supports |
| 3 | Trauma–Depression Hybrid | Trauma screening and regulation-focused therapy | Invest in trauma-informed, cross-domain programs |
| 4 | Trauma–Impulse/Anxiety Blend | Address emotion dysregulation and impulsivity | Build bridge services across trauma and anxiety clinics |
| 5 | ADHD / Conduct / Developmental | Behavioral and developmental interventions | Expand school-based behavioral-health and youth outreach |

Taken together, these implications highlight the practical value of data-driven classification systems in transforming how psychiatric care is organized and delivered. The reproducible, interpretable nature of the six-class structure provides a scalable template for aligning research analytics with real-world clinical triage and policy planning, thereby bridging the gap between computational modeling and applied mental-health reform.

### 5.5 Limitations and Future Directions

While the study provides a strong empirical foundation, several limitations warrant consideration:

**Data Source:** The MHCLD is administrative and may contain diagnostic inconsistencies across states and providers. Future work should validate findings using multi-source or EHR-linked data.

**Binary Indicators:** Modeling disorders as binary simplifies computation but omits severity gradients. Incorporating continuous measures would enhance dimensional fidelity and alignment with HiTOP or RDoC.

**Socioeconomic and Contextual Variables:** The dataset lacks SES, trauma exposure, and service-access data; integrating such variables could reveal structural determinants of comorbidity.

**Cross-Sectional Design:** The study captures a single time point. Future longitudinal modeling (e.g., latent transition or dynamic networks) could illuminate **temporal evolution** of comorbidity.

**Generalizability:** Results reflect treatment-seeking individuals; replication in community and nonclinical samples is needed.

**Methodological Extensions:** Integrating multimodal data (behavioral, biological, genetic) using multilevel mixture or Bayesian network methods could further enhance mechanistic insight.

Addressing these gaps will enable a transition from descriptive to **predictive, mechanism-informed classification systems** that more effectively guide care and policy.

### 5.6 Concluding Remarks

This study demonstrates that psychiatric comorbidity in the U.S. population follows a structured, hierarchical pattern. By integrating LCA, network modeling, and DSM-5 mapping, we identified six transdiagnostic subtypes that reconcile categorical and dimensional models.

The convergence of person-centered and network approaches supports a nested system of psychopathology, wherein individual-level profiles emerge from broader inter-disorder relationships. This hierarchical organization aligns with frameworks like HiTOP and supports a network-informed nosology emphasizing shared mechanisms over isolated diagnoses.

Validated across multiple statistical paradigms, these findings illustrate that complex psychiatric structures can be robustly captured using open, population-scale data. The resulting typology provides a foundation for precision mental-health systems—linking analytic discovery to targeted intervention and equitable policy design.

Ultimately, this research advances a data-driven paradigm for psychiatry, uniting analytic rigor with clinical relevance and offering a scalable roadmap for future transdiagnostic classification and care innovation.

### 5.7 Summary of Research Questions and Hypotheses

This study was guided by five core research questions and corresponding hypotheses, each aimed at uncovering different layers of the comorbidity architecture within the 2021 MHCLD dataset. The table below summarizes how each question was addressed and whether its associated hypothesis was supported by the results.

| RQ | Hypothesis | Key Findings | Supported? |
| --- | --- | --- | --- |
| RQ1: What latent structure best represents the joint occurrence of mental-health disorders within the U.S. treatment-seeking population? | H1: A finite, reproducible set of latent classes will emerge, each corresponding to broad transdiagnostic spectra such as internalizing, externalizing, and psychotic-affective domains (Kotov et al., 2021). | A six-class solution was identified with excellent entropy (0.9999) and high posterior certainty, reflecting interpretable dimensions aligned with broad psychopathology spectra. | Yes |
| RQ2: How do the disorder communities uncovered through network analysis correspond to the individual-level subtypes identified via LCA? | H2: The disorder co-occurrence network will exhibit stable modular clusters that partially align with the LCA-derived classes, reflecting a hierarchical, multi-level structure of comorbidity (Borsboom, 2017; Forbes et al., 2021). | Louvain modularity identified three stable communities; partial overlap with latent classes observed (ARI = 0.33, NMI = 0.51), consistent with hierarchical structure. | Yes |
| RQ3: To what extent do these empirical latent classes parallel the diagnostic groupings defined in *DSM-5*? | H3: Some classes will predominantly align with single *DSM-5* domains (e.g., depressive or psychotic), whereas others will span several domains, revealing transdiagnostic hybrid subtypes that transcend categorical boundaries (McElroy & Borsboom, 2023). | DSM-5 mapping revealed both class-domain concordance (e.g., bipolar–psychotic classes) and cross-domain hybrids (e.g., trauma–depression), supporting both alignment and transdiagnostic patterns. | Yes |
| RQ4: What demographic and clinical patterns distinguish individuals across the latent classes? | H4: Sociodemographic and clinical factors—such as gender, age, and SMI/SED status—will vary systematically across classes in ways that mirror established epidemiological patterns (Kessler et al., 2005; Alegría et al., 2010). | Significant associations observed (Cramér's $V = 0.22–0.33$), with clear patterns by gender, age group, and clinical severity across classes. | Yes |

## 6. Data Availability Statement

The data analyzed in this study are derived from the 2021 U.S. Mental Health Client-Level Data (MHCLD), a publicly accessible dataset curated by the Substance Abuse and Mental Health Services Administration (SAMHSA). The dataset is fully de-identified and available for secondary research use without restriction through the official SAMHSA open-data portal: https://datafiles.samhsa.gov/

All scripts used for preprocessing, modeling, and visualization are available in a public GitHub repository: https://github.com/Supriyabommi/MHCLD-Comorbidity-Analysis-

All scripts, analytic workflows, and processed outputs (CSV, JSON, and PNG files) are available in supplementary files.

## 7. Ethics Statement

This research utilized fully de-identified, publicly available data and did not involve any direct interaction or intervention with human participants.

Accordingly, the study was deemed exempt from institutional review board (IRB) oversight under U.S. federal regulation 45 CFR 46 §101(b)(4), which applies to research involving existing, publicly available datasets. All analyses adhered to the principles outlined in the Declaration of Helsinki (2013) and followed relevant U.S. ethical and data-protection guidelines governing secondary data use.

## 8. Competing Interests

The author declares no commercial, financial, or personal conflicts of interest that could be construed as influencing the research, authorship, or publication of this work.

## 9. Funding

This study did not receive any external funding. All research activities, data analysis, and manuscript preparation were conducted independently.

## Notes

### Competing Interest Statement

The authors have declared no competing interest.

### Author Declarations

Substance Abuse and Mental Health Services Administration, https://www.samhsa.gov/data/

